# Possible association of nucleobindin-1 protein with depressive disorder in patients with HIV infection

**DOI:** 10.1101/2022.07.08.22277410

**Authors:** Yun Yang, Qian Zhang, Jing Yang, Yun Wang, Ke Zhuang, Changcheng Zhao

## Abstract

**Objective:** To investigate the prevalence for depressive disorder among HIV-infected population and preliminarily explore the underlying mechanisms.

**Methods:** Individuals who were newly HIV diagnosed were assessed on the Hospital Anxiety and Depression scale (HAD-A and HAD-D). Then SHIV-infected rhesus monkey model was used to investigate the possible involvement of NUCB1 and CB1 protein in depression-like behavior.

**Results:** The prevalence rate of depression disorder among newly confirmed HIV cases was 27.33% (41/150). The mechanism research results showed elevated NUCB1 levels in cerebrospinal fluid (CSF) from HIV-infected patients suffering from depression were confirmed by western blotting compared to those of HIV-infected patients. Also, immunohistochemical analysis indicated expression of NUCB1 in the cerebral cortex neurons of SHIV-infected monkey was higher than that of healthy control. Conversely, CB1 expression were down-regulated at protein level.

**Conclusions:** Depression are common in HIV infection and associate with NUCB1 expression increase, and NUCB1 may be a potential target for depression among HIV-infected subjects.

## Introduction

Depression is common in human immunodeficiency virus (HIV) infection ^[1][2]^. Studies showed that HIV is a direct and indirect causes of depression. In recent years, the researches that explore possible mechanisms underlying depression among people living with HIV/AIDS (PLWHA), thereby discovering new diagnostic markers and novel therapeutic targets, have become hotspots in the field of both AIDS and neurobiology research.

HIV can enter central nervous system. A paper indicated that an adult-viable mutant that completely disrupts the G protein α-subunit binding and activating motif of nucleobindin 1 (NUCB1) ^[3]^, has been shown to play a neuroprotective role in *Drosophila* model of neurodegenerative disease. Besides, in 2020, a LC-MS/MS analysis predicted that a significantly positive correlation between plasma protein levels of NUCB1 and degree of depression ^[4]^. Therefore, we put forward the hypotheses that alterations in NUCB1 expression might be linked to depression among PLWHA. The present study aimed to survey the prevalence for depression disorder among PLWHA and preliminarily explore the underlying mechanisms. Considering simian immunodeficiency virus (SIV) or simian / human immunodeficiency virus (SHIV) infected rhesus macaques (RMs) have been widely utilized in pathogenic mechanisms of neuroAIDS ^[5][6]^, to unravel the possible involvement of NUCB1 in the pathophysiology of depression among PLWHA, SHIV infection in RMs induces virus replication and affects pathological changes in the brain that resembles that of humans. Understanding of the mechanism underlying the interaction between NUCB1 expression and depression in the context of HIV-infection may provide insights that may facilitate the development of biomarker for diagnosis, new drug target, and treatment response.

## Material and Methods

### Research subjects

In order to eliminate the potential effects of other factors, from January 2015 to October 2020, a total of 150 HIV/AIDS individuals newly confirmed by positive HIV viral load, positive antibody testing (ELISA and western blot), were selected at the First Affiliated Hospital of the University of Science and Technology of China (USTC). All participants were asked to participate in The Hospital Anxiety and Depression (HAD) scale questionnaires and the medical history systems. This study has been reviewed and approved by the Bioethics and Biological Safety Review Committee of USTC (2021-N(H)-236).

### Investigation Procedures

HAD scale was used to assess severity of anxiety and depression symptoms, where A represents anxiety, while D represents depression. The score of each subscale ranges between 0 and 21 points. Overall, the total scores of HAD-A and HAD-D were classified as normal (0 - 7) and anxiety or depressive (8 - 21) with higher scores indicating higher levels of symptoms.

### Western Blot Assay

Night cerebrospinal fluid (CSF) samples (HAD-D score >= 16) were obtained by lumbar puncture in the morning. 3 mL CSF was taken from each subject. Western blot was performed as previously described ^[7]^. Briefly, Proteins in 500 µl CSF sample were separated by SDS-PAGE and transferred onto nitrocellulose membranes. After blocking with 5% milk for 1 h, membranes were incubated with primary antibodies (anti-NUCB1 Rabbit pAb, ABclonal, A3994) at 4°C overnight. Appropriate secondary antibodies (1:2000, Santa Cruz) were used for two-hour incubation at room temperature. Membranes were visualized by ECL plus kit (GE Healthcare and Life Science).

### Animals and Ethics Statement

The RMs used in this study were from the Tianqin Breeding and Research Center (SCXK (e) 2021-0010), Hubei Province, China. The monkeys were housed in an air-conditioned room with an ambient temperature of 16-26°C, a relative humidity of 40-70% and a 12-hour light-dark cycle at the Animal Bio-Safety Level-III laboratory of the Wuhan University (SCXK (e) 2019-0013). The RMs were individually housed in stainless steel wire-bottomed cages with sufficient space (800 mm wide, 800 mm depth and 1600 mm height) and provided with a commercial monkey diet. In addition to animal health was monitored daily by the animal care staff and veterinary personnel. All study protocols were approved by the Institutional Animal Care and Use Committee of USTC (2021-N(A) -349) in accordance with the regulations of the National Institute of Health “Guide for the Care and Use of Laboratory Animals” and all details of animal welfare and steps taken to ameliorate suffering were in accordance with the recommendations of the Weatherall report, “The use of nonhuman primates in research”.

### Virus inoculation and sample collection

Seven Chinese-origin RMs from three different projects were enrolled in this study. Of these, five RMs were inoculated with 10^3^-10^8^ TCID_50_ of SIV/SHIV by either intravenous or intravaginal route under anesthesia with intramuscular injection of ketamine hydrochloride (10 mg/kg) plus intramuscular injection of atropine (0.04 mg/kg), while two healthy RMs were used as negative controls.

### Histology and Immunohistochemistry

At day of sacrifice, a depressive monkey and a healthy control were anesthetized with ketamine-HCl and euthanized by intravenous pentobarbital overdose. Formalin-fixed, paraffin-embedded brain sections from the cerebrum had been obtained, 4 microns sections were processed and stained with hematoxylin and eosin (H&E) staining and immunohistochemistry (IHC). For IHC, sections were done using an automated system, the Dako Autostainer Link. Formalin-fixed paraffin sections were rehydrated with water. Heat-induced epitope retrieval was performed with the FLEX TRS High-pH Retrieval Buffer for 20 minutes. After peroxidase blocking, the specific monoclonal antibody (IHC-plus CNR1/CB1 pAb, Lifespan, LS-B8253; anti-NUCB1 Rabbit pAb, ABclonal, A3994) was applied at room temperature for 20 minutes. The FLEX + Rabbit EnVision System was used for detection. DAB chromogen was then applied for 10 minutes. Slides were counterstained with Mayers hematoxylin for 5 seconds and then dehydrated and coverslipped. Images were then processed and analyzed using CaseViewer software (2.1v, 3D Histech Ltd, Budapest, Hungary). Negative controls were included in the run.

### Western blotting

Western blotting was performed in cerebral cortex tissue lysates, as previously described. Briefly, CD8þ cells from SHIV-infected RMs were lysed with radioimmune precipitation assay buffer. Subsequently, proteins were transferred onto nitrocellulose membranes (Bio-Rad), and appropriate primary antibodies and HRP-conjugated secondary antibodies were used, and proteins were detected with the enhanced chemiluminescent (ECL) reagent (Thermo Scientific), followed by quantification using ImageJ software.

### Statistical Analysis

Statistical analysis of the data was performed using chi-square test (level of significance was 0.05) with SPSS software version 23.0 and GraphPad prism 5.0. In each aspect, univariate analysis was used to determine the variables significantly related to the dependent variable. The confidence interval was 95%.

## Results

### Prevalence of depression among HIV-infected patients

Among 150 participants newly testing positive for HIV, 6.00 % (9/150) of patients met the criteria for a case of depression (HAD-D score >=8) and the prevalence of baseline anxiety was 14.00 % (21/150) according to the HAD-A score >=8 criterion, and combined anxiety and depression accounts for 21.33% (32/150) of the variance in reported body dissatisfaction. Furthermore, the number of HIV-infected individuals with HAD-A score >= 8 was reduced significantly from 53 at baseline to 27 at week 8 (*p* = 0.006), but no statistically significant differences in the number of patients with HAD-D score >= 8 was observed comparing baseline to week 8 (*p* = 0.189, two sample t-test) (See Table 1).

**Table 1.** Prevalence of anxiety and depression among HIV/AIDS patients.

| Items | HAD-A |  | HAD-D |  |
| --- | --- | --- | --- | --- |
|  | Week 0 | Week 8 | Week 0 | Week 8 |
|  | ≤7 | ≥8 | ≤7 | ≥8 |
| Score ≤ 7 | 97 | 105 | 109 | 105 |
| Score ≥ 8 | 53 | 27 | 41 | 27 |
| $\chi^2$ | | 7.443 | | 1.723 |
| *p* |  | 0.006 |  | 0.189 |

### Univariate analyses of variables related to symptoms of depression among individuals with HIV infection

We included personal information and clinical information in univariate analyses to observe whether the variables were related to anxiety and depressive symptoms. As shown in Table 2, among the several items, “age” and “marital status” were significantly related to the depression in those patients (both *p* < 0.05). And, “weight” was significantly related to the patient’s anxiety symptom (*p* = 0.043) among the variables.

**Table 2.**
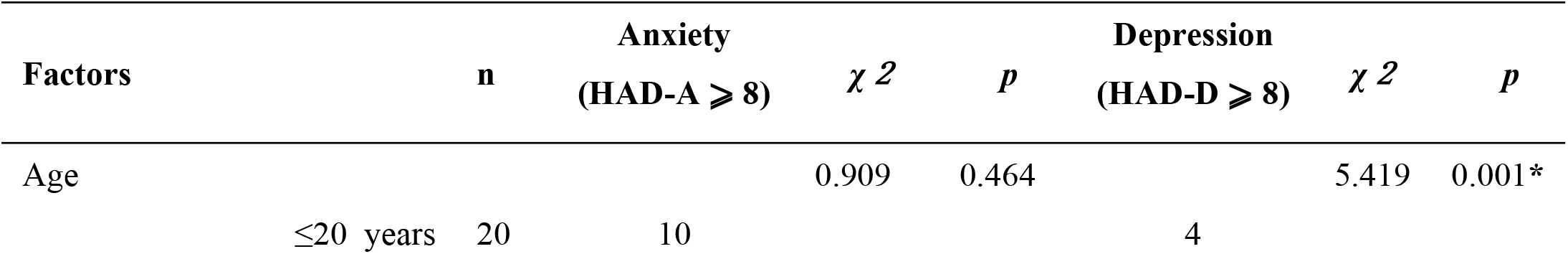
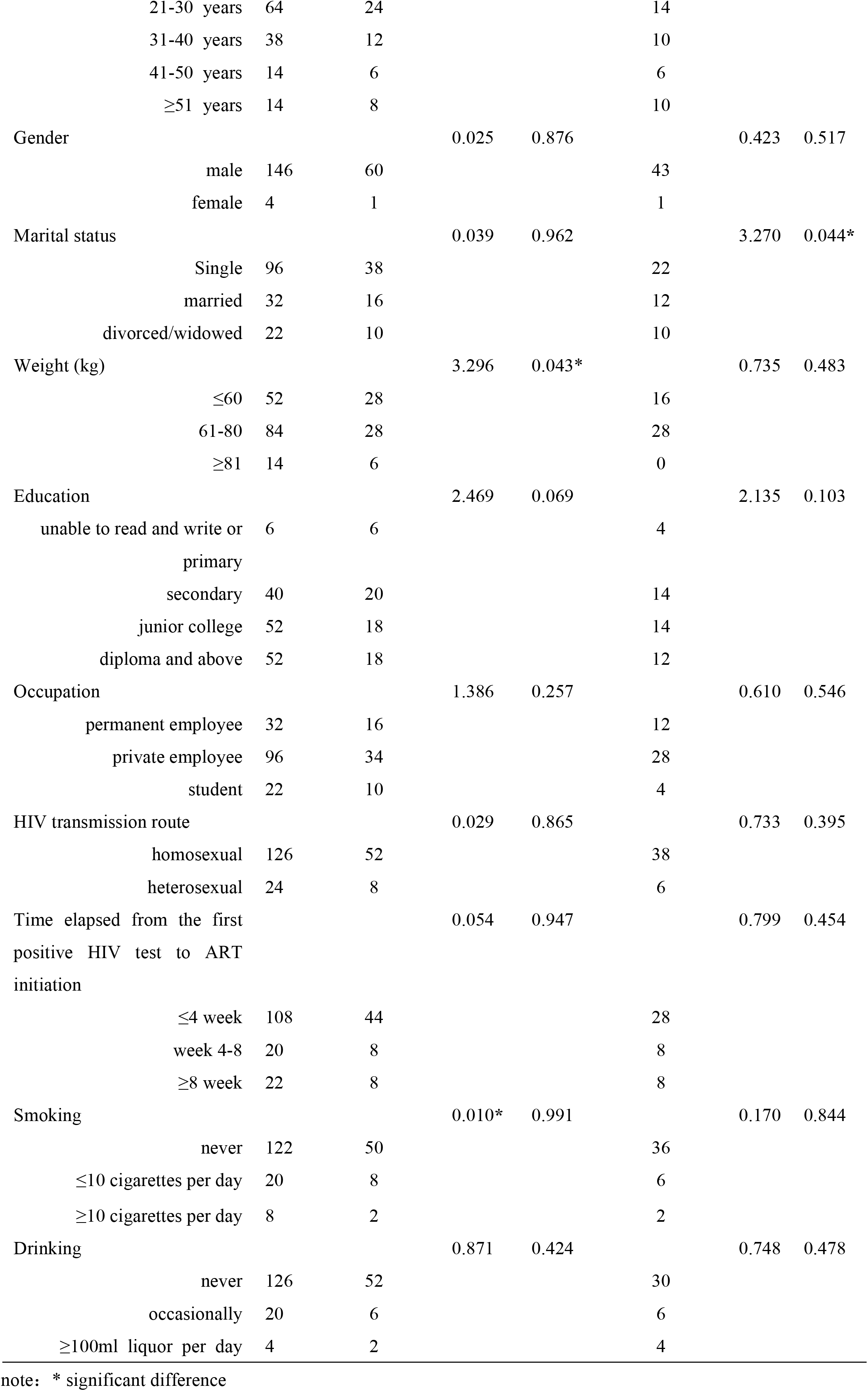
Factors associated with anxiety and depression among patients with HIV/AIDS.

### NUCB1 levels are elevated in CSF from HIV cases currently suffering from depression

CSF is a proximal fluid which communicates closely with brain tissue and contains numerous brain-derived proteins. Thus NUCB1 expressions of CSF in HIV-infected individuals were examined via western blot analysis. As shown in Figure 1, comparing with only HIV cases, the protein expression of NUCB1 in the CSF was significantly increased in those HIV cases currently suffering from depression. Notably, we did not enroll a control group of healthy subjects because of the invasiveness of the CSF procedure for which the Ethics Committee did not allowed.

**Figure 1.**
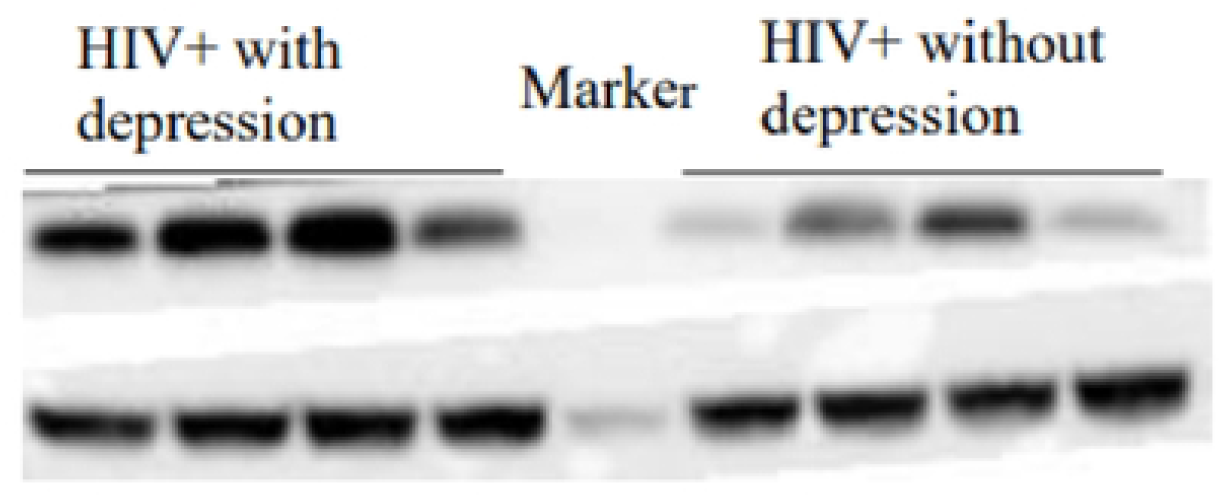
Representative image of protein levels of NUCB1. Western blotting results showed that in HIV-infected individuals, having comorbid depression significantly increased the expression of NUCB1 in the cerebrospinal fluid.

### SHIVKU-1 infection triggers a reduction in the number of neurons in cerebral cortex of rhesus monkey

To explore the pathogenesis of depression among PLWHA, in this study, SIV/SHIV of five RMs established persistent infections. Of note, during the late stage of infection, Macaque WSH01 infected with SHIV_KU-1_ presented depression-like syndromes that mimic those observed in human neuroAIDS, including difficulties in standing, head tilting, weakening of muscle strength, decreased appetite and movement, loss of body mass, ataxia, fear, psychomotor changes, sleep disturbance, and total loss of motoric function on the left side of the body. The animal had a slow progressing course lasting for about 18 months after the triggering infection and was euthanized at week 72. This study demonstrated that SHIV_KU-1_ infected RMs can resemble human neuroAIDS and will become an important tool for studying pathogenesis and evaluating treatment and preventive drugs of neuroAIDS.

As shown in Figure 2, H&E staining showed marked histological damages with increased infiltration and vasculitis in the brain of Macaque WSH01. Importantly, the cerebral cortex exhibited a decrease in the number of neurons. Accompanied with purulent meningitis, the intactness of cerebral cortex could be not revealed more clearly and many cavities appeared after liquefaction of cerebral cortex tissue (Figure 2B). Specifically, the neuronal loss in the cerebral cortex of the SHIV_KU-1_-infected monkey was observed and also substantial changes in the spatial arrangement of neurons. In addition, residual nerve cells bodies in the cerebral cortex changed from swelling to shrinkage, Nissl body staining was weak, the dendritic length due to cell death or an inflammatory process was irregular, and neural axone was thin (Figure 2b).

**Figure 2.**
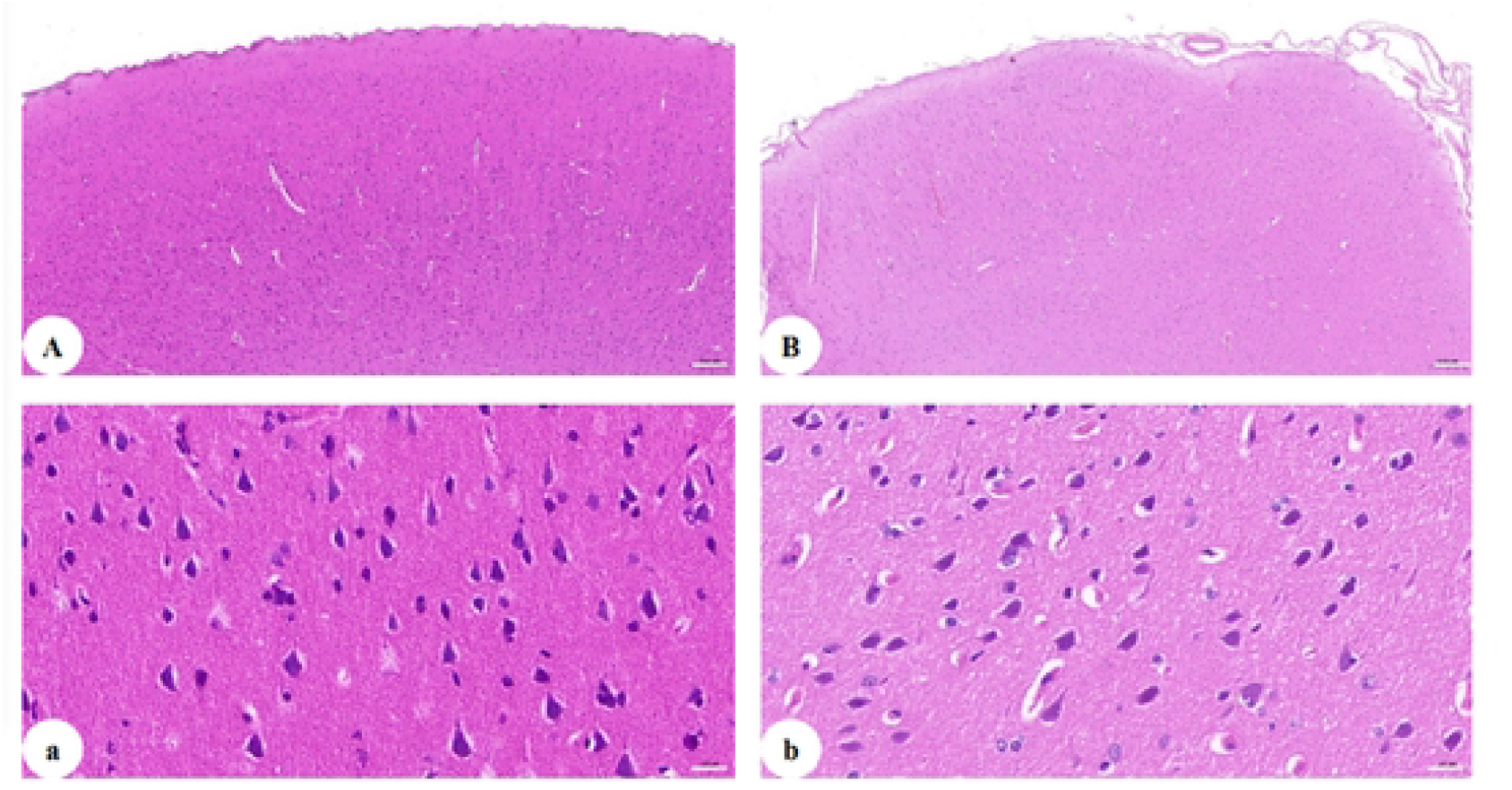
H&E staining of the cerebral cortex. A. section of the cerebral cortex of healthy monkey (50 ×); B. section of the cerebral cortex of the SHIV_KU-1_-infected monkey brain (50 ×); a. section of the cerebral cortex of healthy monkey (400 ×); b. section of the cerebral cortex of the SHIV_KU-1_-infected monkey brain (400 ×).

### Upregulation of NUCB1 protein in cerebral cortex of SHIV_KU-1_-infected monkey

We next verified whether alterations in the protein expression of NUCB1 occur in SHIV_KU-1_- infected monkey via IHC staining analysis. The results revealed that, NUCB1 protein was expressed in cerebral cortex of healthy monkey but, in the affected, the expression of NUCB1 protein was higher (Figure 3). Furthermore, these findings were supported by data from quantitative Western blotting of NUCB1 protein levels in whole cerebrum in vivo (*p* = 0.0056). The results suggest that NUCB1 plays an important role in the pathological processes leading to depression.

**Figure 3.**
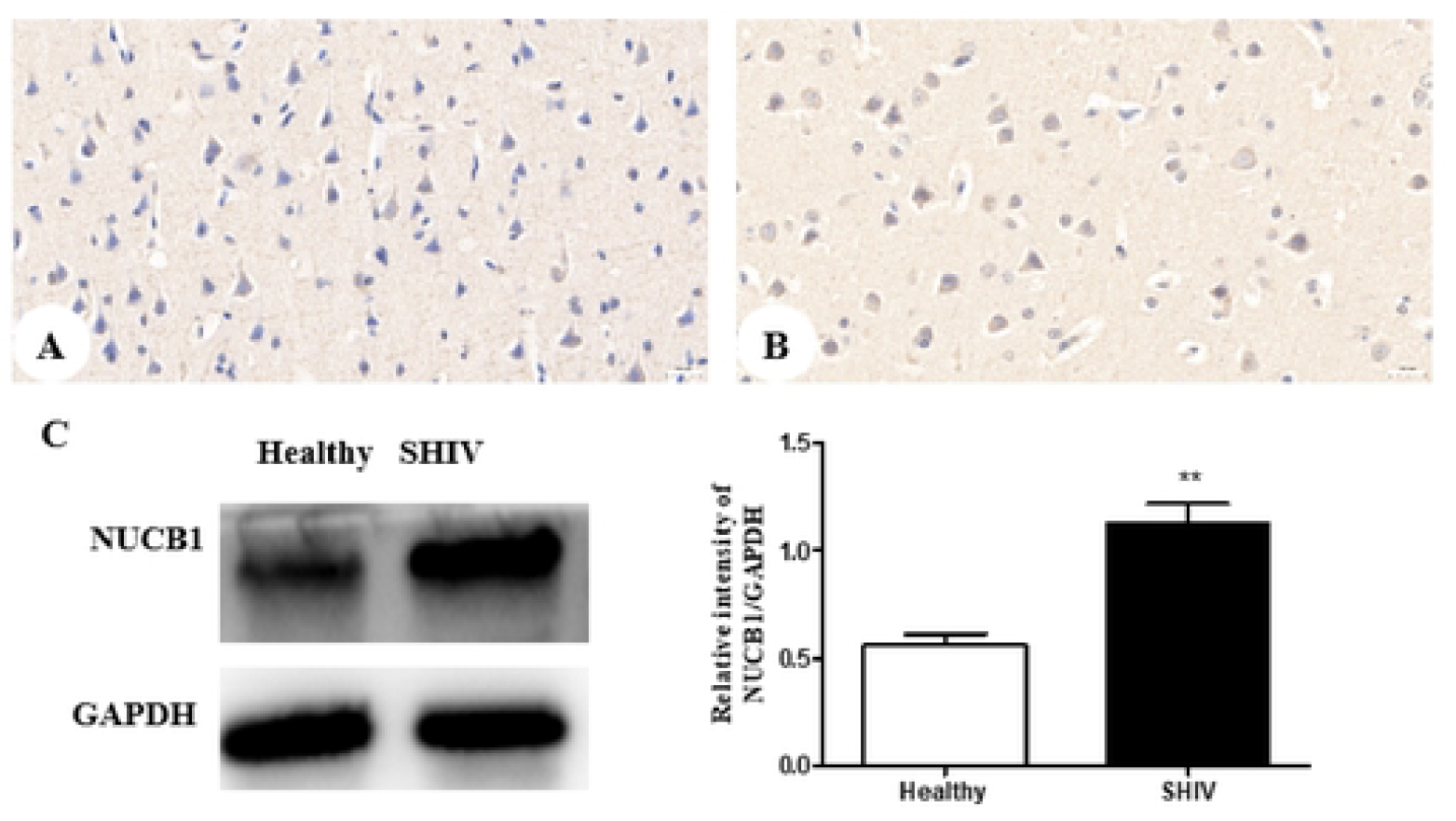
NUCB1 protein expression in cerebral cortex. A. the healthy control (400 ×); B. SHIV_KU-1_-infected monkey (400 ×). C. Comparison of NUCB1 expression of cerebral cortex in healthy control and SHIV_KU-1_-infected monkey using Western blotting. In the column, from *t* test, statistically significant differences of two groups can be determined (*p* = 0.0056).

### Downregulation of CNR1 protein in cerebral cortex of SHIV_KU-1_-infected monkey

Finally, 16 potential targets relative to depression were acquired from the TCMSP database (https://old.tcmsp-e.com/disease.php?qd=228), and further analyzed by the online STRING database (http://string.embl.de/) to explore the functional mechanism of NUCB1 protein.

The network showed CNR1, neuroprotective and highly expressed in the neurons ^[8][9]^, is in close contact with NUCB1 (Figure 4A). Thereby, we investigated the impact of SHIV_KU-1_ infection on CNR1 expression in neurons. Accordingly, we observed, using immunohistochemical technique, the downregulation of CNR1 protein expression in the infected monkey compared to the healthy control (Figure 4B and 4C). This was supported by protein levels of CNR1 via western blot analysis (*p* = 0.0366) (Figure 4D). We speculate that decreased CNR1 expression by SHIV_KU-1_ infection results in depressive disorder, because there is support for existing cannabinoid signaling pathways that can decrease neuronal injury ^[10]^.

**Figure 4.**
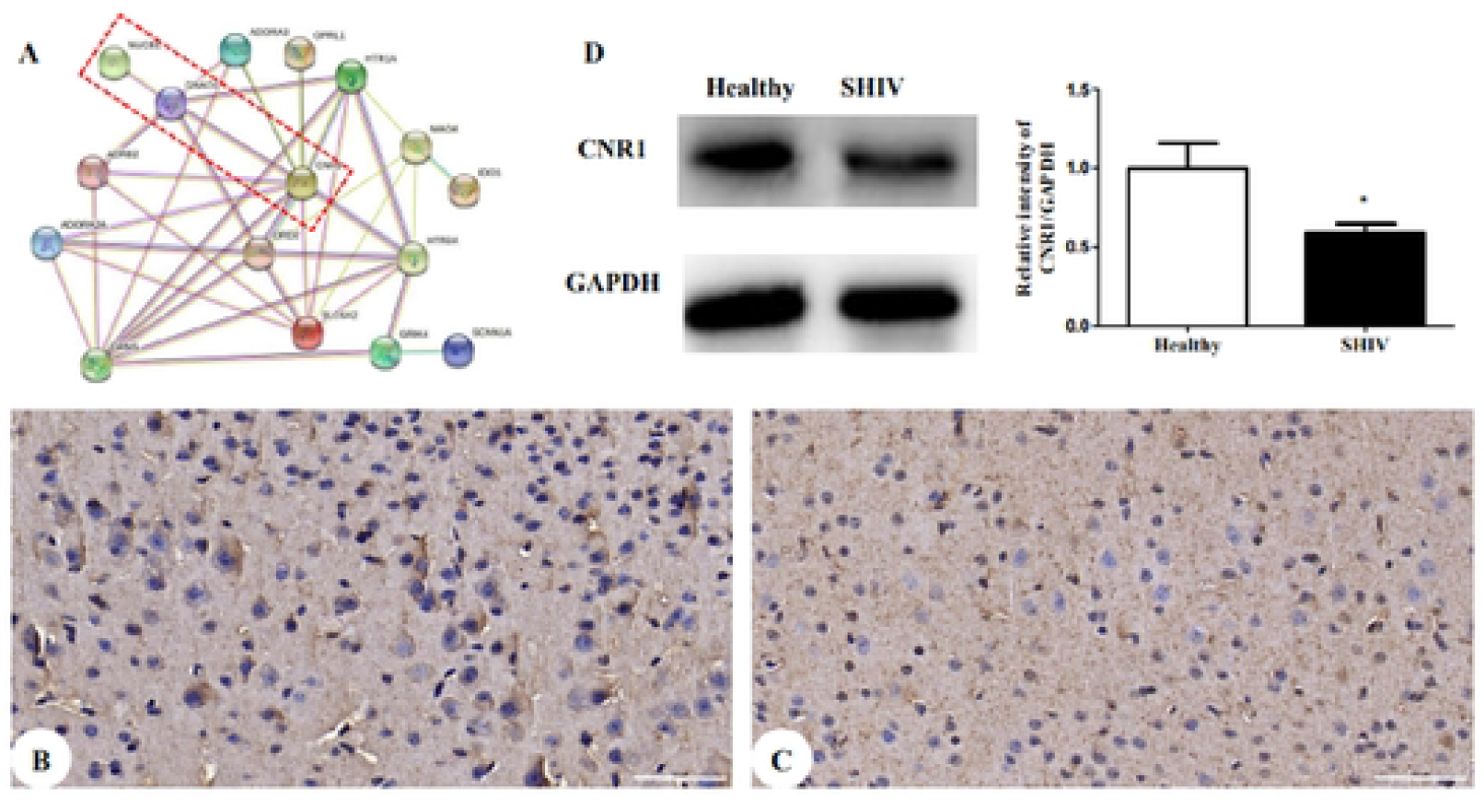
CNR1 protein expression in both SHIV_KU-1_-infected monkey and healthy control. A. The protein-protein interactome networks. Blue rectangle nodes represent downregulated proteins. Red circular nodes stand for the upregulated proteins. The lines represent the regulation of relationship between two nodes. B. IHC staining against CNR1 of the healthy control (400 ×); C. IHC staining against CNR1 of SHIV_KU-1_-infected monkey (400 ×). D. Comparison of CNR1 expression of cerebral cortex in healthy control and SHIV_KU-1_-infected monkey using Western blotting. In the column, from *t* test, statistically significant differences of two groups can be determined (*p* = 0.0366).

## Discussion

Many studies have reported significantly higher prevalence of depression in HIV-infected patients when compared to general population. In the present study the prevalence of depression among HIV-infected patients was 27.33% (41/150). Moreover, univariable logistic regression analysis indicated that age and marital status were associated with depression (*p* < 0.05). Unfortunately, the current diagnoses of depression are still based on clinical manifestations and self-rating scales as the main diagnostic criteria, as a lack of relevant objective laboratory indicators. Thus, there is an urgent need to search for and identify new clinical biomarkers of depression.

Delightfully, our understanding of the (endocannabinoid system, ECS) comprised neuromodulatory lipids and their receptors associated with depression has increased ^[11][12]^. CNR1, also known as CB1, is the most abundant G protein-coupled receptor (GPCR) in the mammalian brain. It was reported that Rocha *et al*. ^[13]^ found that CNR1 knockout mice decreased weight and appetitive processes, reduced rearing and exploratory behaviors, and increased anxiety, compared to wild-type littermates. These symptoms was similar to human depression patients. In addition, a clinical phase I/II trial with SR14716A (rimonabant), a CNR1 antagonist agonist showed that it produced serious neuropsychiatric adverse events such as anxiety, depression, and even suicidal ideation ^[14][15]^. NUCB1, also known as CALNUC or NUC, is a highly conserved, multifunctional protein widely expressed in nervous cells ^[16]^. Herein, down-regulation of CNR1 expression and up-regulation NUCB1 expression in neurons were found in the co-occurrence of depression disorder and SHIV infection, as detected by IHC and Western blot. Several studies has been showed that HIV gp120 stimulates increased cortical fatty acid amide hydrolase (FAAH), subsequently allows for rapid 2-AG and AEA production ^[17]^, selective ligands for the NUCB1 linked to degree of depressive behavior ^[18]^, in postsynaptic neuron. Whereas the decreased CNR1 expression promote the release of 2-AG and AEA ^[19]^. Based on our experimental research and the published data, we propose that a schematic presentation of a possible mechanism of NUCB1 involvement in depression in PLWH (Figure 5). A published article indicated that ZiBuPiYin recipe, which was recorded in the book of Bujuji written by Cheng Wu in the Qing dynasty, prevents and treats diabetes-associated cognitive decline by regulating the NUCB1 protein level ^[20]^. Whether the recipe, through the similar molecular pathways, relieves depressive disorder in PLWHA remains to be further investigated. There are several limitations in our study. Firstly, further studies are needed to demonstrate the expression of NUCB1 in both selective brain areas and specific neuronal subpopulations. Also, animal model, unable to conduct questionnaires, interviews, or oral reports, cannot be quantified the severities of depression. Third, there is no way for a human investigator to really know whether a monkey is feeling depressed. What we can do is observe the behavioral that a monkey makes to viral infection. Finally, there’s too little data on NUCB1 levels in CSF from HIV-infected individuals currently suffering from depression, to draw a convincing conclusion, the sample size should be enlarged, etc. Despite all this, this study suggest that NUCB1 protein may be a novel clinical biomarker for depression, and inhibiting its activity might have potential function that predict and monitor responsiveness of treatment.

**Figure 5.**
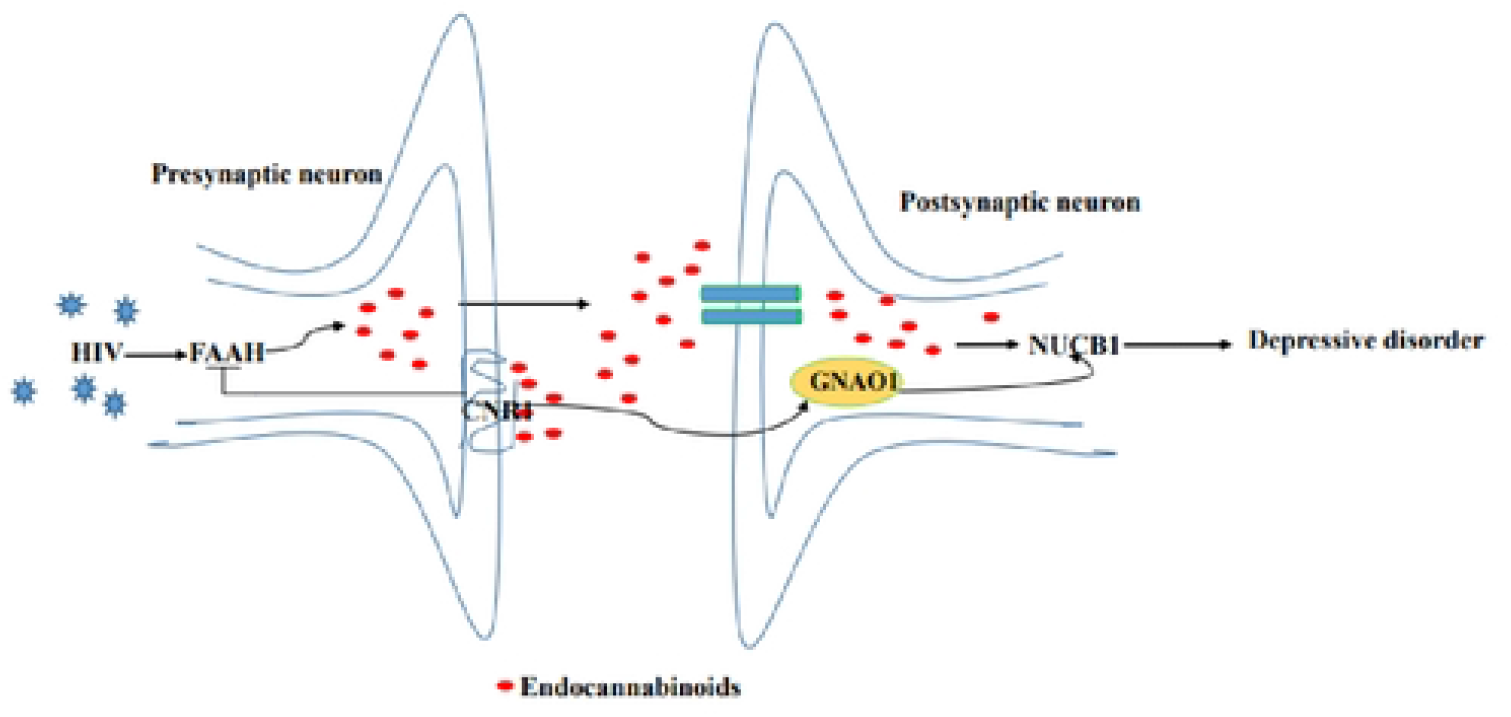
Proposed model of the molecular events in NUCB1-mediated HIV comorbid depression.

## Data Availability

the data are all contained within the manuscript and/or Supporting Information files, enter the following: All relevant data are within the manuscript and its Supporting Information files.

## Acknowledgments

We thank Ying Dong, Fei Li and Ting Liu for their constant support and useful discussions.

## Informed consent

Informed consent was obtained from all individual participants included in the study.

## References

1. Mills JC, Harman JS, Cook RL, et al. Comparative effectiveness of dual-action versus single-action antidepressants for the treatment of depression in people living with HIV/AIDS. J Affect Disord. 2017;215:179–186.

2. Algoodkar S, Kidangazhiathmana A, Rejani PP, et al. Prevalence and Factors associated with Depression among Clinically Stable People Living with HIV/AIDS on Antiretroviral Therapy. Indian J Psychol Med. 2017;39(6):789–793.

3. Balasubramanian V, Srinivasan B. Genetic analyses uncover pleiotropic compensatory roles for Drosophila Nucleobindin-1 in inositol trisphosphate-mediated intracellular calcium homeostasis. Genome. 2020;63(2):61–90.

4. Kim EY, Ahn HS, Lee MY, et al. An Exploratory Pilot Study with Plasma Protein Signatures Associated with Response of Patients with Depression to Antidepressant Treatment for 10 Weeks. Biomedicines. 2020;8(11):455.

5. Omeragic A, Kayode O, Hoque MT, et al. Potential pharmacological approaches for the treatment of HIV-1 associated neurocognitive disorders. Fluids Barriers CNS. 2020;17(1):42.

6. Williams K, Westmoreland S, Greco J, et al. Magnetic resonance spectroscopy reveals that activated monocytes contribute to neuronal injury in SIV neuroAIDS. J Clin Invest. 2005 Sep;115(9):2534–45.

7. Yu J, Li X, Matei N, et al. Ezetimibe, a NPC1L1 inhibitor, attenuates neuronal apoptosis through AMPK dependent autophagy activation after MCAO in rats. Exp Neurol, 2018, 307, 12–23.

8. Ma L, Jia J, Niu W, et al. Mitochondrial CB1 receptor is involved in ACEA-induced protective effects on neurons and mitochondrial functions. Sci Rep. 2015;5:12440.

9. Schurman LD, Lichtman AH. Endocannabinoids: A Promising Impact for Traumatic Brain Injury. Front Pharmacol. 2017;8:69.

10. Liu Q, Bhat M, Bowen WD, et al. Signaling pathways from cannabinoid receptor-1 activation to inhibition of N-methyl-D-aspartic acid mediated calcium influx and neurotoxicity in dorsal root ganglion neurons. J Pharmacol Exp Ther. 2009;331:1062–1070.

11. Stampanoni Bassi M, Gilio L, Maffei P, et al. Exploiting the Multifaceted Effects of Cannabinoids on Mood to Boost Their Therapeutic Use Against Anxiety and Depression. Front Mol Neurosci. 2018;11:424.

12. Rana T, Behl T, Sehgal A, et al. Integrating Endocannabinoid Signalling In Depression. J Mol Neurosci. 2021;71(10):2022–2034.

13. Rocha L, Cinar R, Guevara-Guzmán R, et al. Endocannabinoid System and Cannabinoid 1 Receptors in Patients With Pharmacoresistant Temporal Lobe Epilepsy and Comorbid Mood Disorders. Front Behav Neurosci. 2020;14:52.

14. Ettaro R, Laudermilk L, Clark SD, et al. Behavioral assessment of rimonabant under acute and chronic conditions. Behav Brain Res. 2020;390:112697.

15. Nguyen T, Thomas BF, Zhang Y. Overcoming the Psychiatric Side Effects of the Cannabinoid CB1 Receptor Antagonists: Current Approaches for Therapeutics Development. Curr Top Med Chem. 2019;19(16):1418–1435.

16. Kamthan A, Kamthan M, Kumar A, et al. A calmodulin like EF hand protein positively regulates oxalate decarboxylase expression by interacting with E-box elements of the promoter. Sci Rep. 2015;5:14578.

17. Towe SL, Meade CS, Cloak CC, et al. Reciprocal Influences of HIV and Cannabinoids on the Brain and Cognitive Function. J Neuroimmune Pharmacol. 2020;15(4):765–779.

18. Niphakis MJ, Lum KM, Cognetta AB 3rd, et al. A Global Map of Lipid-Binding Proteins and Their Ligandability in Cells. Cell. 2015;161(7):1668–80.

19. Hermes DJ, Yadav-Samudrala BJ, Xu C, et al. GPR18 drives FAAH inhibition-induced neuroprotection against HIV-1 Tat-induced neurodegeneration. Exp Neurol. 2021l;341:113699.

20. Xu H, Zhou W, Zhan L, et al. The ZiBuPiYin recipe regulates proteomic alterations in brain mitochondria-associated ER membranes caused by chronic psychological stress exposure: Implications for cognitive decline in Zucker diabetic fatty rats. Aging (Albany NY). 2020;12(23):23698–23726.

